# Mini-XT, a miniaturized tagmentation-based protocol for efficient sequencing of SARS-CoV-2

**DOI:** 10.1101/2021.09.29.21263685

**Authors:** Marc Fuchs, Clara Radulescu, Miao Tang, Arun Mahesh, Deborah Lavin, Syed Umbreen, James McKenna, Mark Smyth, Eilís McColgan, Zoltan Molnar, Chris Baxter, Timofey Skvortsov, Aditi Singh, Fiona Rogan, Julia Miskelly, Stephen Bridgett, Derek Fairley, David A. Simpson

## Abstract

**Introduction:** The COVID-19 pandemic has highlighted the importance of whole genome sequencing (WGS) of SARS-CoV-2 to inform public health policy. By enabling definition of lineages it facilitates tracking of the global spread of the virus. The evolution of new variants can be monitored and knowledge of specific mutations provides insights into the mechanisms through which the virus increases transmissibility or evades immunity. To date almost one million SARS-CoV-2 genomes have been sequenced by members of the COVID-19 Genomics UK (COG-UK) Consortium. To achieve similar feats in a more cost-effective and sustainable manner in future, improved high throughput virus sequencing protocols are required. We have therefore developed a miniaturized library preparation protocol with drastically reduced consumable use and costs.

**Methods:** SARS-CoV-2 RNA was amplified using the ARTIC nCov-2019 multiplex RT-PCR protocol and purified using a conventional liquid handling system. Acoustic liquid transfer (Echo 525) was employed to reduce reaction volumes and the number of tips required for a Nextera XT library preparation. Sequencing was performed on an Illumina MiSeq.

**Results:** We present the ‘Mini-XT’ miniaturized tagmentation-based library preparation protocol available on protocols.io (https://dx.doi.org/10.17504/protocols.io.bvntn5en). The final version of Mini-XT has been used to sequence 4,384 SARS-CoV-2 samples from N. Ireland with a COG-UK QC pass rate of 97.4%. Sequencing quality was comparable and lineage calling consistent for replicate samples processed with full volume Nextera DNA Flex (333 samples) or using nanopore technology (20 samples). SNP calling between Mini-XT and these technologies was consistent and sequences from replicate samples paired together in maximum likelihood phylogenetic trees.

**Conclusion:** The Mini-XT protocol maintains sequence quality while reducing library preparation reagent volumes 8-fold and halving overall tip usage from sample to sequence to provide concomitant cost savings relative to standard protocols. This will enable more efficient high-throughput sequencing of SARS-CoV-2 isolates and future pathogen WGS.

## Introduction

The COVID-19 pandemic has highlighted the importance of large-scale whole genome sequencing (WGS) of SARS-CoV-2. This enables surveillance of virus epidemiology, tracking of virus transmission and identification of variants with greater infectivity or potential for vaccine escape. This work has been driven in the UK by an integrated national SARS-CoV-2 genomic surveillance network (The Coronavirus Disease 2019 (COVID-19) Genomics UK Consortium) [1, 2].

Many approaches have been developed to enrich SARS-CoV-2 for WGS, including target capture, and virus-specific reverse transcription [3-5]. While these have some advantages, random primed reverse transcription followed by multiplex tiled PCR has been most widely adopted. The ARTIC protocol[6] has been used in many studies [7, 8] and employs two pools of primers, which are being continually improved [9], to amplify ∼400bp tiled amplicons.

Particularly in the early stages of the pandemic nanopore sequencing was used to provide rapid turnaround for single or small batches of samples. Increasing prevalence and recognition of the value of sequencing all positive cases has driven demand for higher throughput methods. Although nanopore protocols now enable multiplexing of 96 samples, sequencing platforms with greater output, such as Illumina, are more suitable.

Both the cost and availability of reagents are critical determinants of the number of samples that can be processed. The disruption caused by the pandemic to manufacturing and the logistics of reagent delivery, in combination with a significantly increased global demand for viral RNA extraction and sequencing, resulted in delivery delays and temporary shortages of laboratory consumables in many countries, impacting not only COVID-19 testing, but other clinical and fundamental microbiology research [10]. To maximise throughput from a given stock of reagents, protocols have been modified to reduce volumes (eg ARTIC V1 to V3) or been specifically designed to minimize the reagents required, such as CoronaHIT [11].

Automation is an essential part of any high throughput protocol, but unfortunately processing of multiple plates requires many expensive plastic pipette tips. This also raises logistical challenges due to COVID-related delays and has longer term sustainability issues. Acoustic liquid handling enables the transfer of low volumes in the nanolitre range without the use of physical tips [12]. We and others have previously applied this technology to the preparation of, for example, synthetic DNA [13], RNA-Seq [14-16] and plasmid libraries. Here we present a ‘Mini-XT’ protocol for sequencing of SARS-CoV-2 using ARTIC tiled amplicons, Nextera XT-based library preparation miniaturized using an Echo 525 liquid handling system (Beckmann) and sequencing on an Illumina MiSeq. Details are available on protocols.io (https://dx.doi.org/10.17504/protocols.io.bvntn5en). This enables a 10-fold reduction in reagents relative to the standard Nextera XT protocol and use of significantly fewer disposable plastic tips. We have used the Mini-XT protocol to generate over 4000 SARS-CoV-2 genome sequences, with quality comparable to those prepared using full volume DNA Flex library preparation or on the nanopore platform.

## Methods

### Samples

A total of 4,384 samples are reported in detail (a further 4,586 have been processed with earlier versions of the Mini-XT protocol (Supplementary Figure 1)). These are Pillar 2 (non-clinical swab testing of general population) swab extracts prepared by Randox Laboratories (Crumlin, Northern Ireland) using the MagMAX Viral/Pathogen Nucleic Acid Isolation Kit (MVP II) (Thermo Fisher Scientific, MA).

### cDNA generation

RT-PCR followed the ARTIC Network nCoV-2019 sequencing protocol v3 (LoCost) (https://www.protocols.io/view/ncov-2019-sequencing-protocol-v3-locost-bh42j8ye) and its modified version adapted for use with Illumina sequencers (https://dx.doi.org/10.17504/protocols.io.bnn7mdhn) [17]. cDNA setup was performed in a pre-PCR environment and bench surfaces and pipettes were disinfected before starting work. SARS-CoV-2 viral nucleic acid extracts and two non-template controls containing nuclease-free water per 96-well plate (NTCs), were reverse transcribed with LunaScript RT SuperMix Kit (NEB), using 5 µl of the RNA sample in a total reaction of 10 µl. The reactions were incubated for 2 minutes at 25°C, followed by 20 min at 55°C and 1 minute at 95°C before cooling to 4°C.

### Amplicon generation

The cDNA samples were amplified by tiled PCR using separate primer pools derived from premixed ARTIC nCoV-2019 V3 panel (100 µM, IDT) diluted 1:10 in molecular grade water, to achieve 10 µM primer stocks. Two PCR reactions per sample plate, for primer Pools A and B, were performed in a total volume of 25 µl using 4 µl of each 10 µM primer set, 5 µl of 5x Q5 Reaction Buffer (NEB), 0.5 µl of dNTPS (10 mM each), 0.5 U of Q5 Hot Start High-Fidelity DNA Polymerase (NEB) and 2.5 µl of cDNA. The cycling conditions were: heat activation at 98°C for 30 seconds followed by 35 cycles of denaturation at 98°C for 15 seconds and annealing at 63°C for 5 min and then a hold at 4°C.

### Amplicon purification and quantification

A pair of PCR A and PCR B plates were centrifuged at 280*g* and 10 µl from each well of plate A and B combined in a fresh fully skirted PCR plate. Automated bead cleaning was performed using a Biomek NXp robot (Beckman Coulter. CA) with an 8 channel head using 1.5x KAPA Pure beads (Roche) to sample ratio. Samples were eluted in 60 µl 10 mM Tris pH 8.0 and quantified using an Invitrogen Quant-iT dsDNA broad range assay kit (Q33130) with a PHERAstar FS multimode microplate reader (BMG Labtech).

### Mini-XT library preparation

The quantified RT-PCR products (A+B amplicons) were normalised to 0.2 ng/µl using the Echo 525 Liquid Handler (Beckman Coulter): four 96-well plates with RT-PCR products were combined into one 384-well Echo source plate (384PP 2.0), whereby each source pate well was filled with 30 µl A+B amplicon. The Echo Liquid Handler was then programmed to transfer a volume of each amplicon mix containing 8 ng of DNA into a second Echo source plate (normalisation plate). Each well of the normalisation plate was manually topped up with 40 µl of 10 mM Tris buffer at pH 8.0, resulting in the desired target concentration of 0.2 ng/µl.

The normalised amplicons were prepared for sequencing using the Nextera XT DNA Library Preparation Kit (Illumina) in combination with the Nextera XT Index Kit v2 Set A and Set D. The manufacturer recommended volumes were scaled down 10-fold and the reactions prepared using the Echo Liquid Handler. For the tagmentation reaction, 1000 nl Tagment DNA Buffer was transferred from a 6-reservoir Echo source plate (001-11101) into a 384-well PCR plate (Library preparation plate), followed by 500 nl of amplicons from the normalisation plate and 500 nl of Amplicon Tagment Mix from a 384PP 2.0 plate. After spin-down, the plates were incubated at 55°C for 5 minutes on a Tetrad DNA Engine 2 thermal cycler (Bio-Rad) and then placed on ice. Five hundred nl of Neutralize Tagment Buffer was added to the Library Preparation plate from a 6-reservoir source plate to stop the tagmentation reaction, followed by a 5 minute incubation at room temperature.

The library amplification reaction was again prepared with the Echo. The Nextera XT Indexes were presented on a 384PP 2.0 plate and transferred to the Library preparation plate so that each well received one of the 384 possible unique index combinations. After that, 1500 nl of Nextera PCR Master Mix was transferred from a 6-reservoir plate into the Library preparation plate. After another spin-down, the libraries were amplified on a Tetrad DNA Engine 2 in 12 PCR cycles as outlined in the Illumina Protocol for Nextera XT Library preparations.

Up to 384 amplified Libraries were pooled without normalisation and purified in two consecutive 1.6x bead cleanups with KAPA Pure beads. Two wash steps per cleanup were performed with 80% ethanol, and the pool was eluted in 10 mM Tris at pH 8.0, quantified with High sensitivity Qubit dsDNA assay (Thermo Fisher Scientific, MA) and analysed with a Tapestation D1000 assay (Agilent, CA) to determine the average fragment size. The molarity was calculated based on the Qubit concentration and the D1000 size, applying an average molecular weight of 660 g/mol per 1 bp of dsDNA:

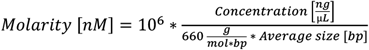

### DNA Flex library preparation

Full volume Illumina DNA Prep library preparation (formerly called Nextera DNA Flex Library Prep), catalogue number 20018705, was performed according to the manufacturer’s protocol, using IDT® for Illumina® DNA/RNA UD Indexes Set A or B, catalogue number 20027213 or 20027214.

### Illumina Sequencing

The Library pool was diluted to 4 nM with 10 mM Tris pH 8.0, denatured with 0.2 N NaOH solution and diluted to 20 pM with Hyb Buffer following Illumina instructions for the Miseq. The 20 pM pool was further diluted to 9 pM and spiked with 1% PhiX at 12.5 pM before being loaded onto a sequencing cartridge. 150 bp paired-end sequencing was performed on a MiSeq system using the 300-cycle v2 MiSeq Reagent Kit. FASTQ generation was performed on-board with the Local Run Manager v3.0, secondary analysis was performed off-board as described below.

### Nanopore library preparation

The Nanopore library preparation protocol was adapted from nCoV-2019 sequencing protocols v2 (https://dx.doi.org/10.17504/protocols.io.bdp7i5rn) and v3(https://protocols.io/view/ncov-2019-sequencing-protocol-v3-locost-bh42j8ye) and the amended protocol is available on protocols.io: https://www.protocols.io/private/8BFC7458DD268B846ADA0B5CA5A71C57. All reagents are detailed in (Supplementary File 1).

NEBNext Ultra II End Repair/dA-Tailing Module (E7546, New England Biolabs, MA) was used for end preparation of purified amplicon, for which 1.2 μl Ultra II End Prep Reaction Buffer and 0.5 μl Ultra II End Prep Enzyme Mix was added to 50ng of purified amplicon and nuclease-free water added to a volume of 10 μl per reaction. The mixture was incubated at room temperature for 15 min and 65 °C for 15 min, followed by incubation on ice for 1 min

For sample multiplexing, ligation of barcodes was then performed using NEBNext^®^ Ultra^™^ II Ligation Module (E7595, New England Biolabs, Ipswich, MA, United States) and Native Barcoding Expansion kits (EXP-NBD104/EXP-NBD114/EXP-NBD196, Oxford Nanopore Technologies, Oxford, UK). The end preparation product from the previous step (0.75 μl) was input into the ligation reaction by adding 5 μl Ultra II Ligation Master Mix, 0.15 μl Ligation Enhancer, 1.25 ul Native barcode, and 2.85 μl Nuclease-free water. The mixture was incubated at room temperature for 20 min and 65 °C for 10 min, followed by incubation on ice for 1 min. Samples were pooled together and purified using KAPA Pure Beads with 0.4x beads-to-sample ratio. The barcoded amplicon pool and beads were incubated at room temperature for 5 min, and the pellet was washed twice in 250 μl Short Fragment Buffer (EXP-SFB001, Oxford Nanopore Technologies, Oxford, UK), followed by one wash in 200 μl 70% ethanol. The barcoded amplicon pool was eluted in 30 μl Elution Buffer (LSK-109, Oxford Nanopore Technologies, Oxford, UK) and quantified using Qubit™ dsDNA HS Assay Kit.

The ligation of adaptors was performed using NEBNext^®^ Quick Ligation Module (E6056, New England Biolabs, Ipswich, MA, United States) and Adapter Mix (AMII) from Native Barcoding Expansion kits. The ligation reaction was performed by adding 10 μl Quick Ligation Reaction Buffer (5X), 5 μl Quick T4 DNA Ligase and 5 μl Adapter Mix (AMII) to the 30 μl barcoded amplicon pool from the previous step and incubating the mixture at room temperature for 20 min. The adaptor-ligated amplicon pool and 50 μl beads were incubated at room temperature for 5 min, the pellet washed twice in 250 μl Short Fragment Buffer and the purified library eluted in 15 ul Elution Buffer and quantified using Qubit™ dsDNA HS Assay Kit.

### Nanopore sequencing

MinION sequencing was performed on R9.4 flow cells (FLO-MIN106D, Oxford Nanopore Technologies, Oxford, UK) following manufacturer’s guidelines. MinION sequencing was controlled using MinKNOW™ software. Up to 15 ng of purified library was input into the loading library by adding 37.5μl Sequencing buffer and 25.5 μl Loading beads (LSK-109, Oxford Nanopore Technologies, Oxford, UK). Elution buffer was added to a volume of 75 μl loading library. The run was monitored with the assistance of ARTIC-nCoV-RAMPART-v1.0.0 (https://artic.network/ncov-2019/ncov2019-using-rampart.html).

### Data analysis

For each Illumina sequencing run, the base-calling and demultiplexing of reads by sample-id, was routinely performed by the “MiSeq Reporter” software [18]. The resulting data was then transferred from the Illumina MiSeq computer to the Queen’s University Kelvin2 HPC compute cluster for analysis. (Optionally, if the sample-sheet required modification, the Illumina “bcl2fastq” [19] or “BCL Convert” [20] pipeline was used to base-call and demultiplex the data).

The “connor-lab/ncov2019-artic-nf” nextflow-based open-source pipeline [21], as used by the COG-UK consortium, was then used with its “illumina” and “profile conda” options, and otherwise default options. Internally, this pipeline uses: “TrimGalore” [22] to trim the adapter sequence from the paired reads; then “bwa-mem” [23] with default options to align the trimmed reads to the Wuhan-Hu-1 reference genome (accession id: MN908947.3); “samtools” to sort the aligned reads in the bam files, then “iVar” [24] to trim the ARTIC amplicons and generate a consensus fasta sequence for each sample. To determine which resulting samples are accepted, a python QC script is used to accept those consensus sequences that have either over 50% of reference bases covered by at least 10 reads, or have a stretch of more than 10 Kbp of sequence without N’s (where N’s are defined as bases with less than 10 reads).

To analyse Nanopore data, the ncov2019-artic nextflow pipeline [21] was used to automate the ARTIC network nCoV-2019 novel coronavirus bioinformatics protocol [25]. Firstly, the Guppy program (available from https://community.nanoporetech.com/downloads) was used for basecalling and demultiplexing of reads, using the “require_barcodes_both_ends” options, and arrangement files nb12 and nb24:

~~~
guppy_barcoder --require_barcodes_both_ends -i run_name -s
output_directory --arrangements_files “barcode_arrs_nb12.cfg
barcode_arrs_nb24.cfg”
~~~

Then the ncov2019-artic pipeline was run with the ‘--nanopolish’ option. The pipeline’s nanopore configuration file (https://github.com/connor-lab/ncov2019-artic-nf/blob/master/conf/nanopore.config) specifies the parameters used for the analysis. As the ARTIC protocol can generate chimeric reads, the artic “guppyplex” program is used to accept reads with length of 400 to 700 bases, then samples with fewer than 10 reads are excluded. Barcodes having fewer than 100 reads are ignored. The minimap2 [26] read aligner is used. To call consequences of a variant on the encoded amino acid, the typing frequency threshold was 0.75 and minimum coverage depth 20.

“Pangolin” [27] and “PangoLEARN” [28] were used to assign each qc-passed sample’s genome consensus sequence to the most likely Pango lineage [29]. The minimap2 pair-wise aligner and the “type_variants” [30] script were used to call specific variants of interest/concern, and the “SnipIt” [31] package was used to plot an image of the variants in each sample compared to the Wuhan reference. Data was collated and summarised using bash and python scripts.

### Phylogenetic analysis

Phylogenetic analysis was performed using the ‘Nextstrain’ collection of tools (nextstrain.cli 3.0.3) [32] to analyse our consensus fasta sequences. The linux-based command line module, specifically the Augur module, was used to perform alignment and infer trees.

## Results

This study analysed primarily Northern Irish ‘Pillar 2’ (swab testing for the wider population) SARS-CoV-2 samples from Randox Laboratories, with a smaller number of ‘Pillar 1’ (NHS swab testing for those with a clinical need, and health and care workers) samples from the Belfast Trust used in comparisons between sequencing protocols.

To facilitate high throughput whole genome sequencing of these samples and to reduce consumable use, a miniaturised version of the Nextera XT (Illumina) library preparation protocol called ‘Mini-XT’ was developed. A graphical overview is provided in Figure 1, with the full protocol available at https://dx.doi.org/10.17504/protocols.io.bvntn5en. A list of reagents is provided in Supplementary File 1. We report here a total of 4,384 samples processed using the Mini-XT protocol of which 4271 (97.4%) passed COG-UK QC. This is comparable with the results of the study by Baker et al., in which the maximum pass rate of 97.6% was reported for higher viral load samples with a known Ct of 32 or lower processed using CoronaHiT-Illumina [11]. The sequencing metrics for the runs organised in date order are shown in Figure 2. An extended figure of all runs completed since January 2021 (Supplementary Figure 1) demonstrates the improved consistency as the final protocol was established. One recent run 210622 failed to reach acceptable QC and was excluded from further analysis. As expected, the percentage of samples passing QC closely mirrored the mean percent per sample of bases covered. In addition to read depth the distribution of reads is critical and probably explains why the correlation between the mean number of aligned reads per sample and percentage of samples passing QC was less significant.

**Figure 1.**
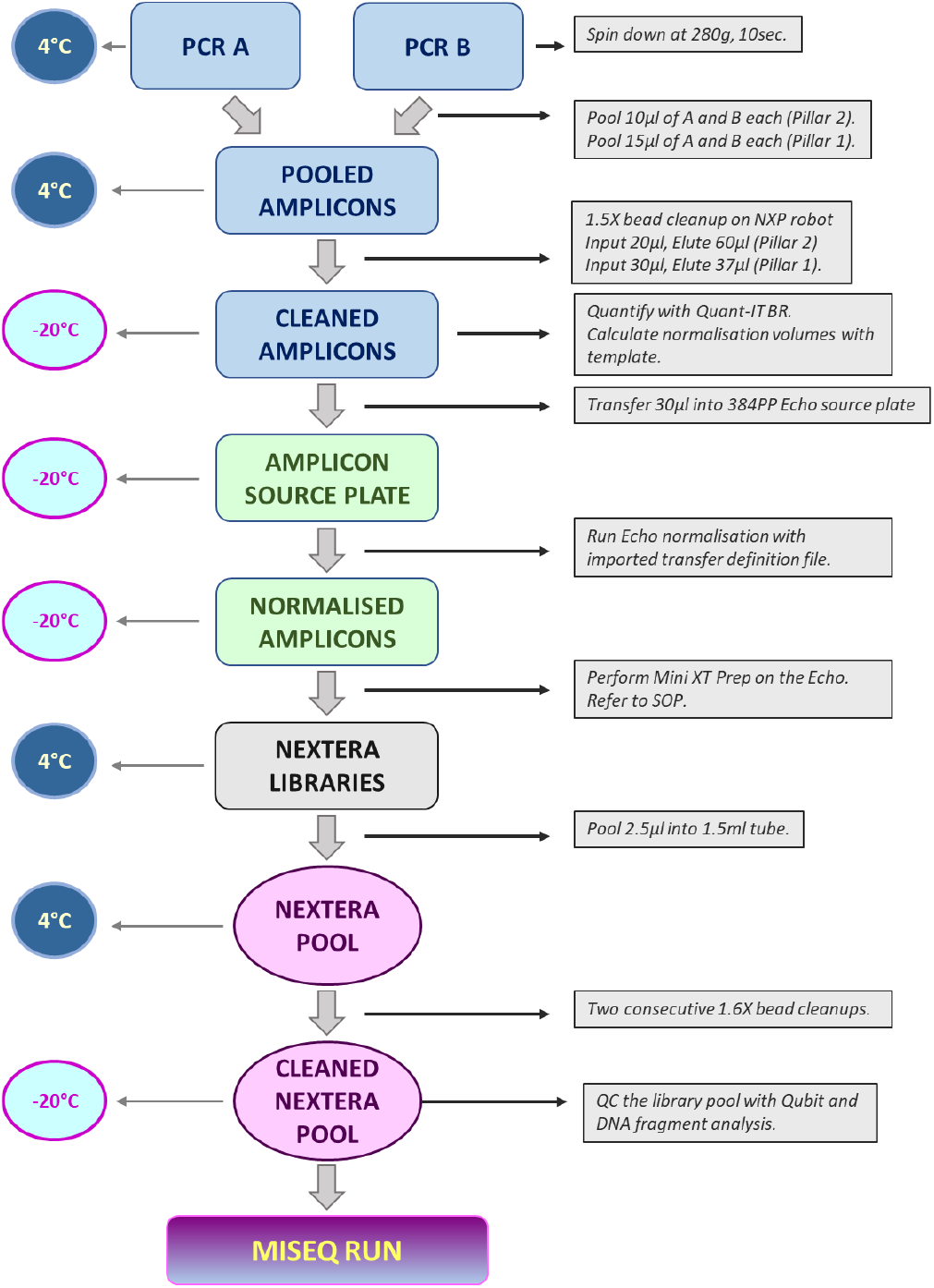
Overview of the Mini-XT protocol. The workflow can be paused at any stage and the plates stored at the indicated temperature.

**Figure 2.**
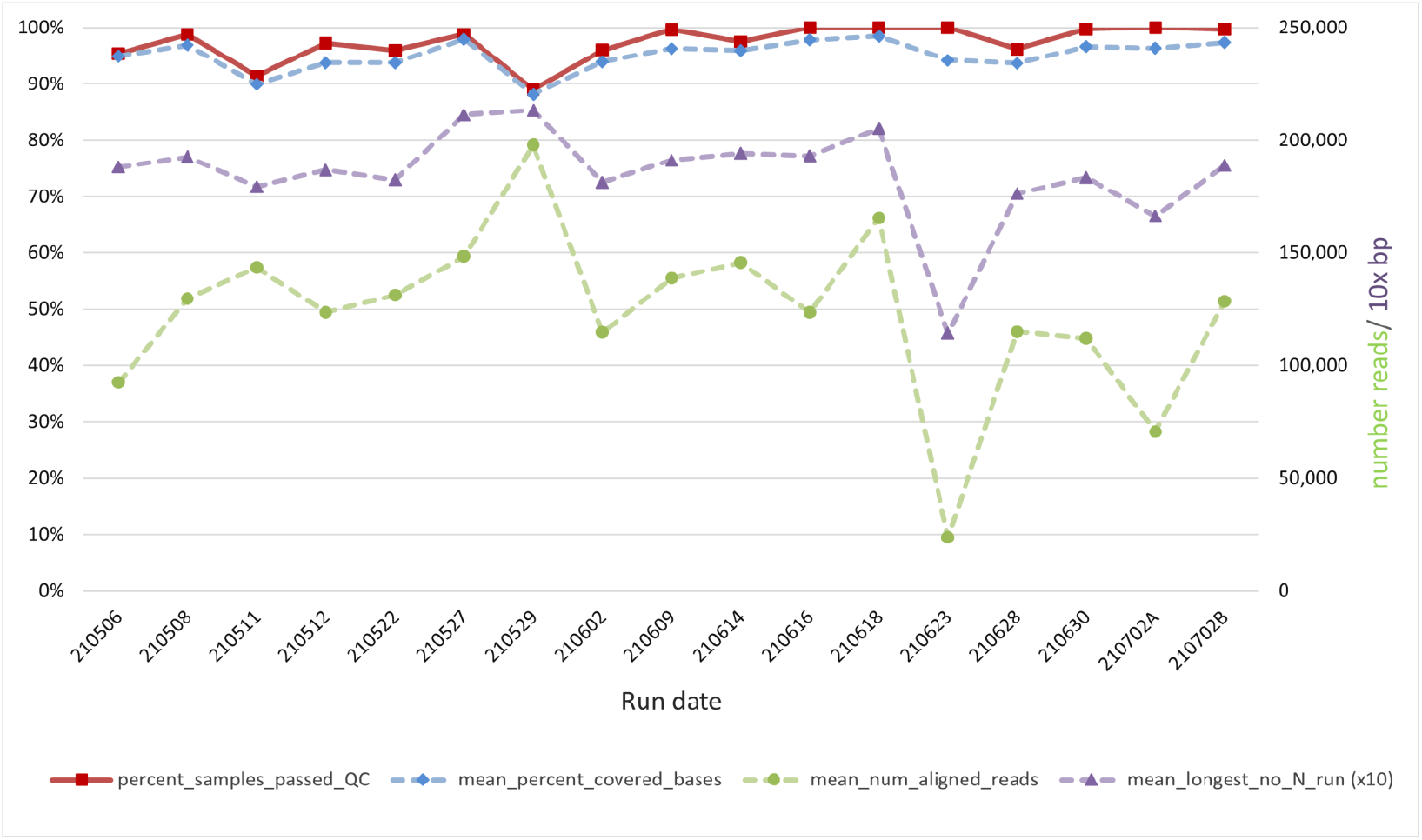
Mini-XT sequencing metrics for individual runs. All runs over a 3-month period are indicated by date (YYMMDD).

As expected, the percent of bases covered by the consensus sequence reduces slightly as the Ct value increases, from over 99% in the most concentrated samples (Ct<20) to 97% in those samples with the highest Ct’s tested (28≤ Ct <30) (Supplementary Figure 2).

To test whether the approximately 10-fold reduction in reagents used in the Mini-XT protocol compromises performance relative to standard full volume DNA-Flex protocols, four 96 well plates of samples were processed in parallel using both approaches. The sequencing metrics, including percent coverage (Figure 3, Supplementary Figure 3) were broadly comparable. For 202 samples the lineage calls from both protocols were identical, for 5 the same but with descendant sublineages missing from one of the calls and for 4 there were discrepancies in sub-lineages between protocols. In these 4 cases most of the characteristic variants are common between the discrepant sublineages (Supplementary Figure 4 [33]) and the differing calls presumably reflect differing sequence quality at the discriminating sites. In a phylogenetic tree created with the consensus sequences generated using Mini-XT and DNA-Flex protocols, the same samples sequenced by different technologies clustered together (Figure 3), reflecting the consistency of SNP calling between methods.

**Figure 3.**
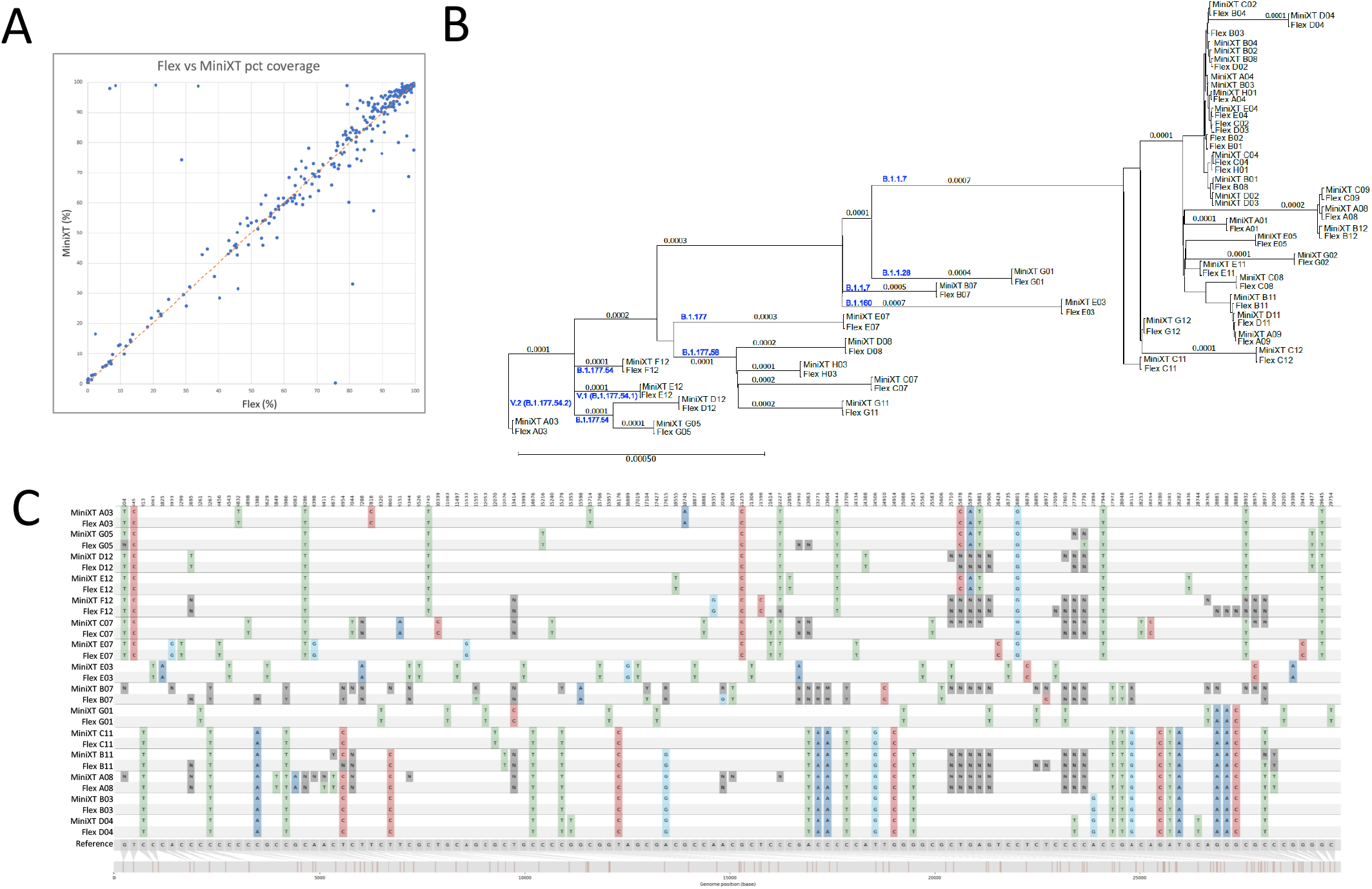
Sequencing results from Mini-XT and Illumina DNA Flex library preparation are comparable. A. The percentage coverage was very similar between each of 333 samples prepared by both library preparation protocols. B. A subset of the consensus sequences (named according to protocol and sample number) were placed in a phylogenetic tree with branch lengths shown in nucleotide substitutions per site and the lineages comprising the main branches indicated. The same samples sequenced using both protocols cluster together on the tree. C. The same SNVs were identified in each sample by both platforms. This is illustrated in a SnipIt plot of 15 pairs of sequences representing each branch of the tree and aligned to a reference Wuhan sequence modified to contain all invariant SNVs identified in these samples.

Nanopore sequencing is widely used throughout the COG-UK consortium [2] and we use this approach for samples requiring a rapid turnaround. To further validate the Mini-XT protocol we compared the results of 20 samples that had also been sequenced using the nanopore platform. The sequencing metrics were broadly comparable (Figure 4A). The lineage calls were identical for the same samples processed using each technology and the sample pairs clustered together when plotted on a phylogenetic tree (Figure 4B).

**Figure 4.**
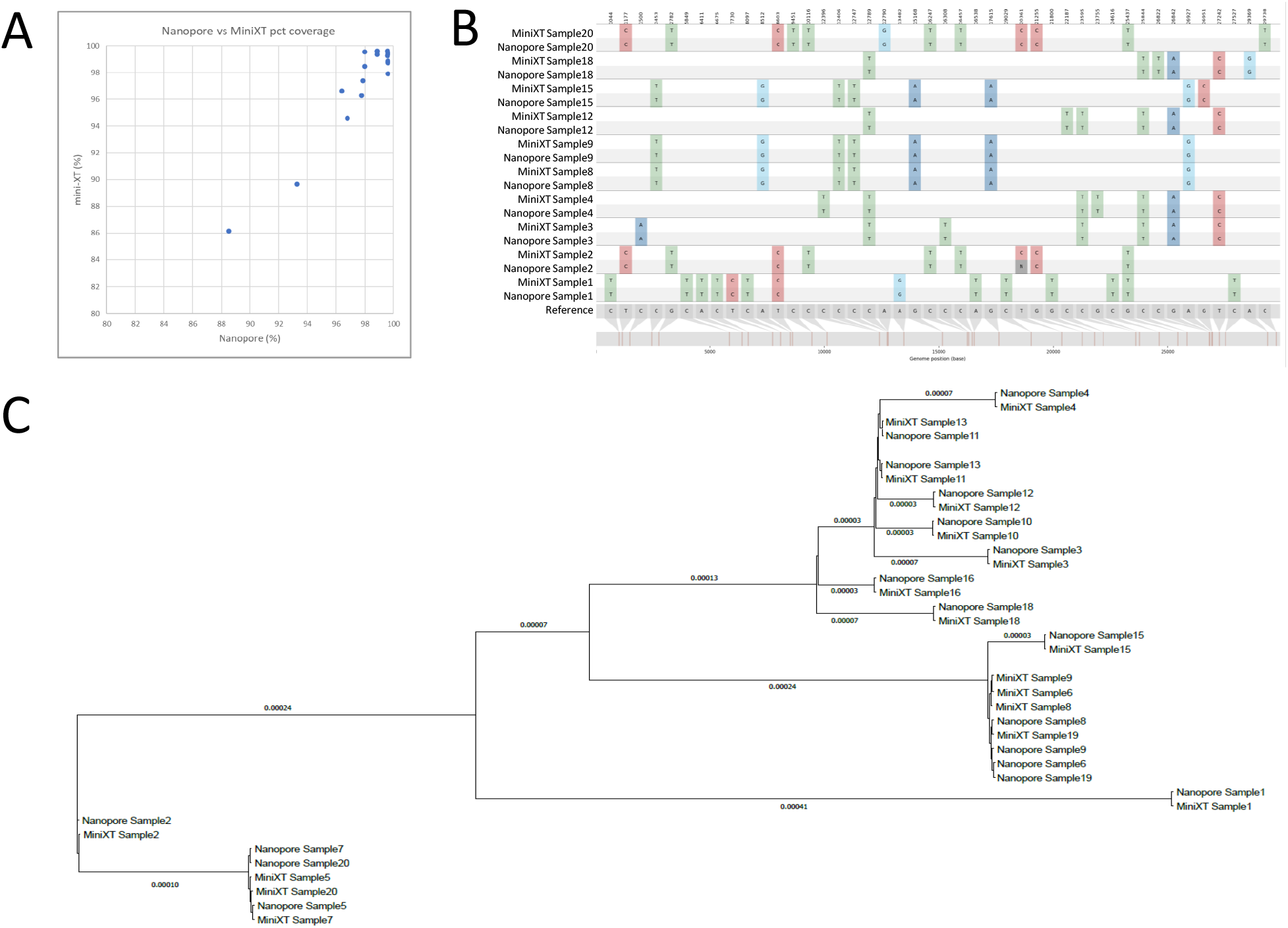
Mini-XT and Illumina sequencing results are comparable with those from Nanopore technology. Twenty samples with B.1.1.7 lineage were sequenced on both platforms. A. The percentage coverage was very similar between platforms. B. The same SNVs were identified by both platforms, as exemplified in a SnipIt plot of 10 pairs of sequences aligned to a reference Wuhan sequence modified to contain all invariant SNVs identified in these samples. C. In a phylogenetic tree of all 20 samples the same samples (named according to protocol and sample number) clustered together, reflecting consistent SNV calling (branch lengths shown in nucleotide substitutions per site).

The use of acoustic liquid transfer enables a significant reduction in the use of consumables, in particular plastic tips. This equates to an approximately 90% reduction in tips required during library preparation and a reduction from 23 to 12 tips per sample for processing from RNA sample to completed library.

New England Biolabs (NEB) have released re-balanced ARTIC primer pools aimed at improving uniformity of SARS-CoV-2 genome coverage and an associated library preparation kit. For a small number of samples tested, the coverage obtained with the ARTIC v3 primers used throughout the rest of this study and the NEB pools was comparable (Supplementary Figure 5). The same amplicon pools processed with Mini-XT or the NEB library preparation protocol produced similar results. A new version of the ARTIC primers (v4) has recently been released. In a comparison between 20 samples amplified with v3 or v4 primers the latest version performed better, with a reduced percent of N bases (0.54% vs 2.17%), increased percent of covered bases (99.4% vs 97.8%) and increased longest no N run (28,919% vs 20,222%) (Figure 5).

**Figure 5.**
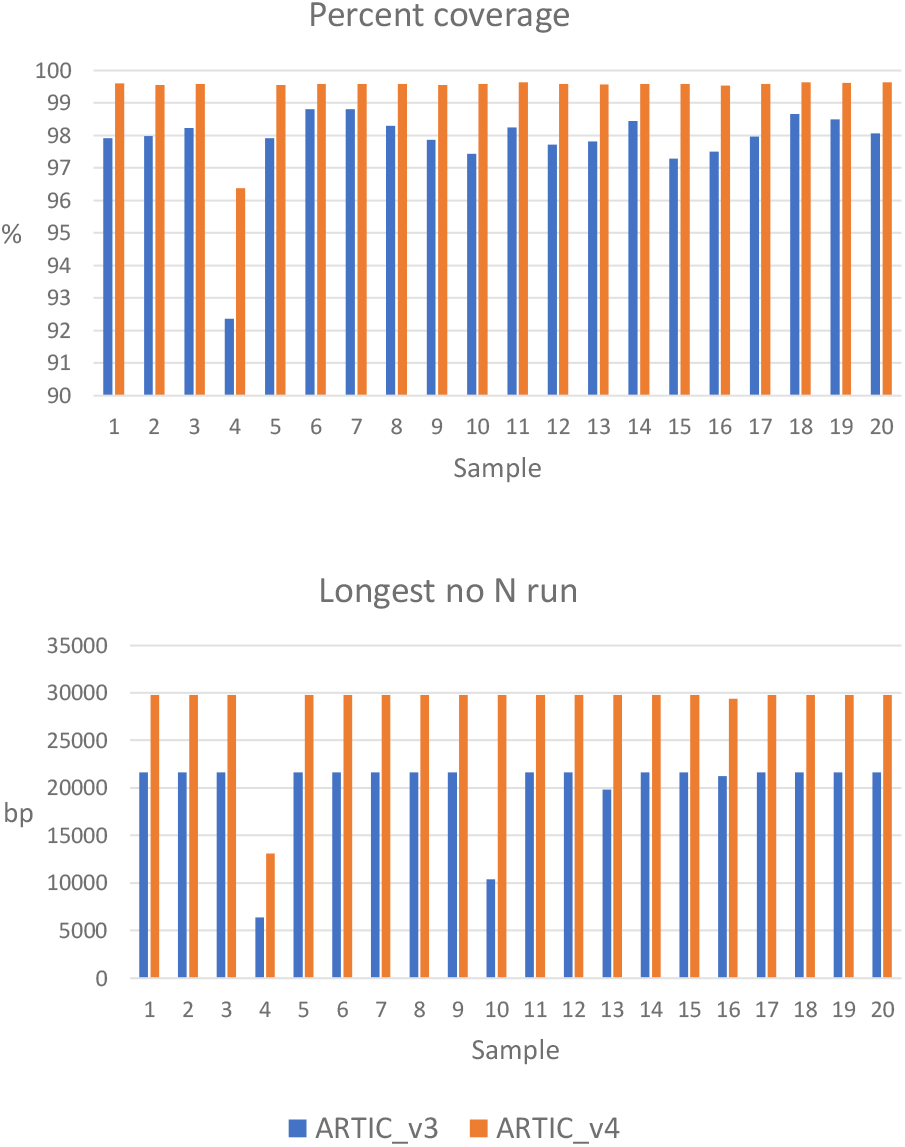
Comparison of samples amplified with ARTIC v3 and v4 primer sets. The percent coverage (top) and longest run with no N’s (bottom) were consistently greater with the v4 primers.

## Discussion

WGS has become a critical part of the international response to the COVID-19 pandemic [1] and will be key in any similar future situations. Indeed, WGS is now an established tool for monitoring many infectious diseases in both the USA [34] and Europe [35]. This increasing demand necessitates the development of more cost-effective high throughput library preparation methods. The ability of the Echo 525 liquid handler to dispense low volumes in a 384 plate format has already been exploited for high throughput low-cost RT-qPCR SARS-CoV-2 surveillance testing [36]. Although various high throughput protocols for sequencing of SARS-CoV-2 have been reported [11, 37, 38], the use of acoustic liquid transfer enabled a greater reduction in volume in the Mini-XT protocol than many alternatives. The ability of the Echo system to dispense variable volumes also facilitated efficient normalization of amplicon pool concentrations. As with other robotic platforms, automation of sample preparation minimises human error and improves consistency.

Similar protocols employing acoustic liquid transfer to reduce reagent volumes for generating libraries for RNA-Seq and synthetic biology applications have been reported previously by us [14] and others [13, 15, 16]. However, Mini-XT is the first such protocol to be applied specifically to whole-genome sequencing of SARS-CoV-2. The confirmation that sequence quality is comparable to that achieved with conventional Illumina or Oxford Nanopore library preparation protocols provides confidence in use of this approach for virus surveillance and research to understand viral transmission and evolution. The thousands of samples processed and reported here demonstrate the robustness of the protocol. While Artic v3 primers were used for amplification of SARS-CoV-2 cDNA throughout the study, the initial findings reported for the v4 primer set suggest that it will improve the quality of sequences generated. The Mini-XT protocol can be used to prepare libraries from any DNA sample, and we anticipate that it will be of use for wider pathogen WGS applications because it can simultaneously decrease costs and dramatically increase sample throughput. Increased use of WGS for public health surveillance and in clinical microbiology are current objectives in Europe [35] that Mini-XT sequencing could facilitate. This approach may also be valuable in the emerging field of clinical metagenomics. Direct metagenome sequencing has the potential to transform diagnostic clinical virology in particular [39, 40]. Wider use of high-throughput sequencing for diagnosis is often limited by cost and capacity constraints, so there is great scope to apply novel methods such as Mini-XT sequencing in this area.

One of the most effective ways for labs to become more sustainable is to reduce single use plasticware [41]. The Mini-XT protocol requires a remarkable ∼4600 fewer tips per 384 samples processed than equivalent conventional manual or robotic preparation. Not only does this reduce waste, but also makes the protocol more resilient to consumable shortages, such as experienced during the COVID-19 pandemic.

Although the Mini-XT protocol is optimized for high-throughput use, which maximizes reagent and plasticware savings to offset capital costs, it can be easily adapted to a 96-sample format.

## Conclusion

We have demonstrated that sequence quality can be maintained after applying acoustic liquid transfer to dramatically reduce reagent volumes during library preparation. The resulting Mini-XT protocol provides an effective, robust, low cost, high-throughput library preparation approach for WGS of SARS-CoV-2. The value of large-scale sequencing of SARS-CoV-2 has been demonstrated during the COVID-19 pandemic and this protocol will facilitate the ongoing provision of whole-genome sequences.

## Data Availability

Routine SARS-CoV-2 genome sequences determined in this study were deposited in the CLIMB database ( https://www.climb.ac.uk/) and are available for download along with all deposited sequences ( https://cog-uk.s3.climb.ac.uk/phylogenetics/latest/cog_all.fasta). Sequences generated in comparative studies are available upon request.

## Acknowledgements

Thanks to the Queen’s University Belfast Genomics CTU (https://www.qub.ac.uk/sites/core-technology-units/Genomics/) for access to facilities and advice. Thanks to staff of the Belfast Health & Social Care Trust Regional Virus Lab (RVL) and Randox Laboratories for provision of positive samples for sequencing. Thanks to Vaughan Purnell for managing the Kelvin2 HPC used for data analysis and configuring storage, cluster job-queues, installing software and giving helpful advice.

## Supplementary files

**Supplementary File 1. Reagents required for the Mini-XT protocol**.

**Supplementary Figure 1.**
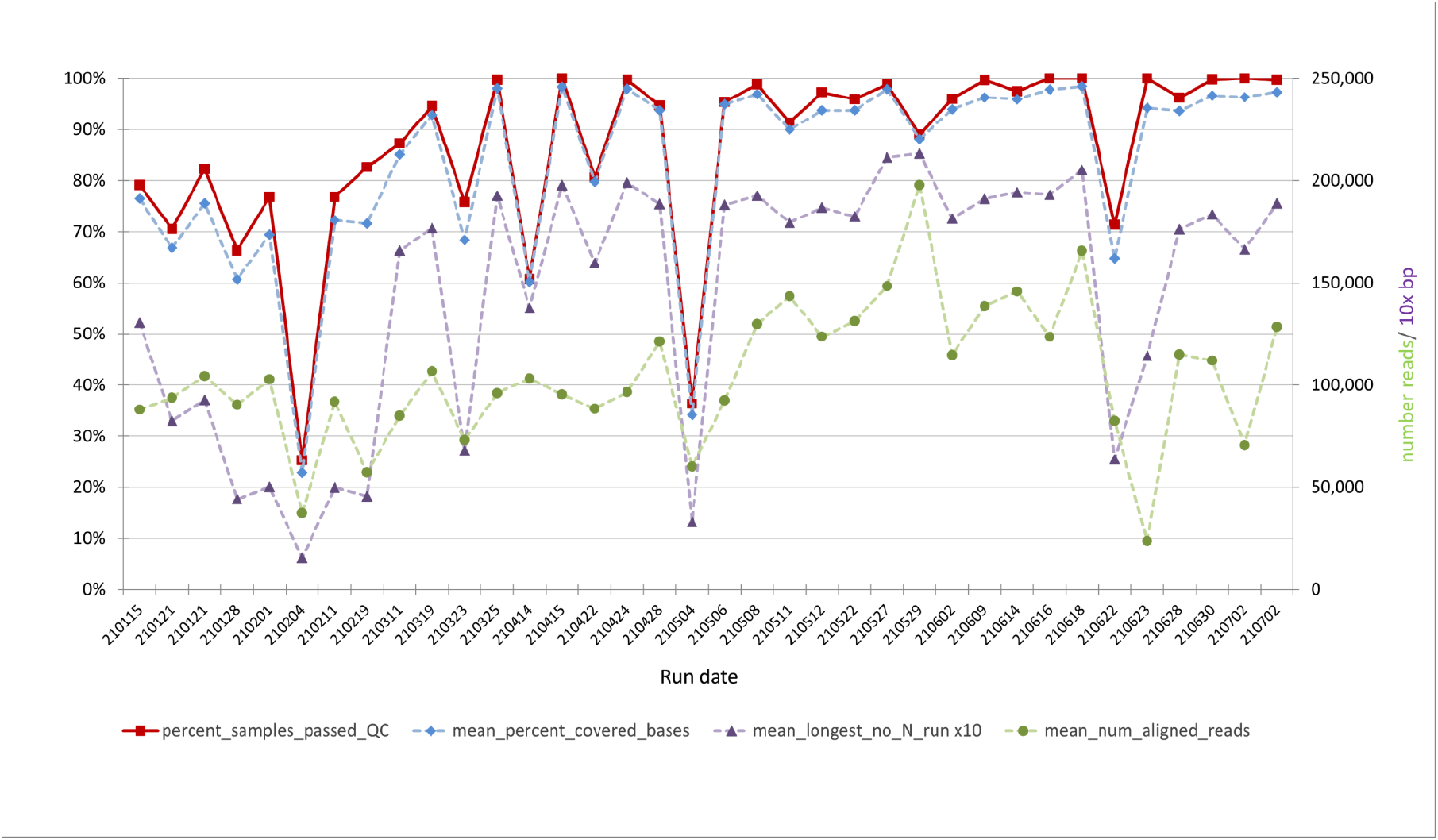
**Mini-XT sequencing metrics for all individual Mini-XT runs indicated by date** (YYMMDD).

**Supplementary Figure 2.**
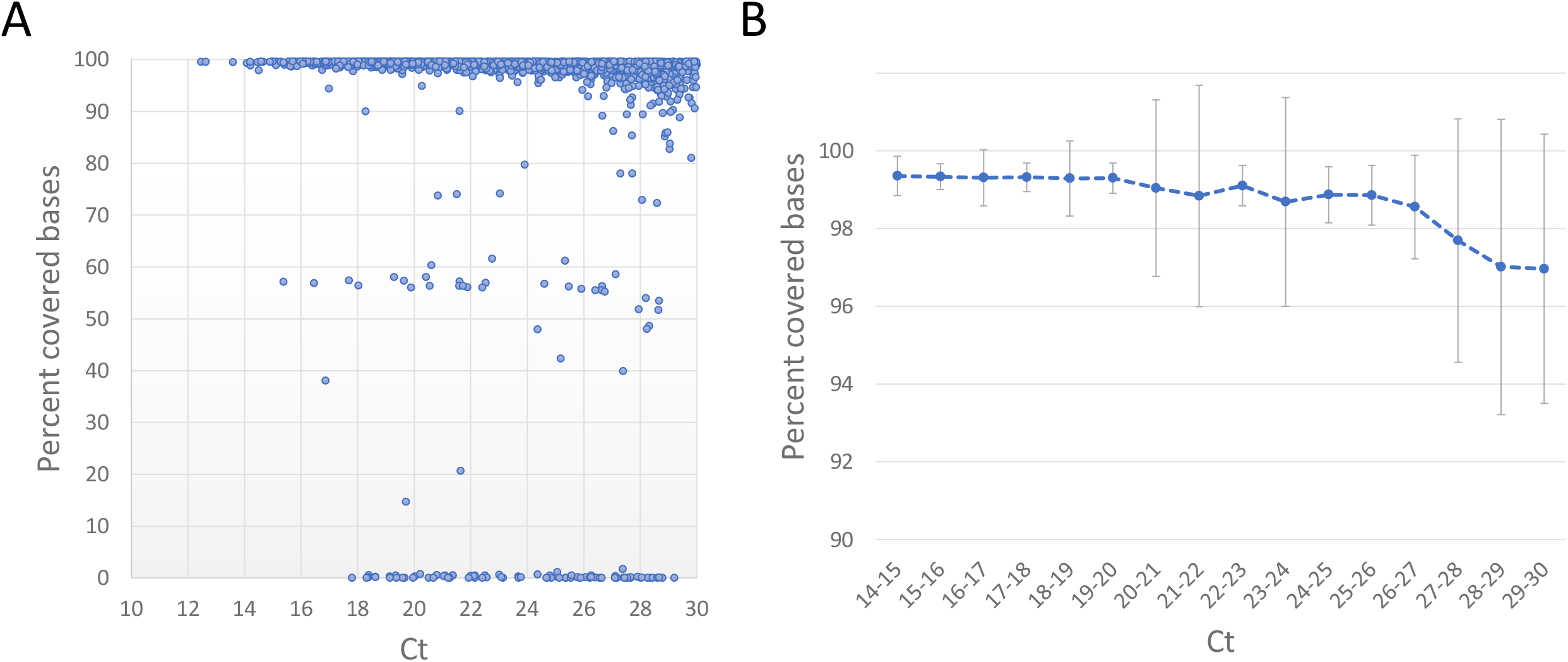
RT-qPCR Ct values of the SARS-CoV-2 positive RNA samples sequenced using Mini-Xt vs percent of bases covered. A. Relationship between Ct value (Orf1ab target gene) and the percent of bases covered in SARS-CoV-2 genome sequences for 2000 samples processed with the Mini-XT protocol. B. Average percent covered bases (+/-SD) for samples with increasing Ct ranges (excluding samples with one or both amplicon pools failing, ie percent covered bases <70)

**Supplementary Figure 3.**
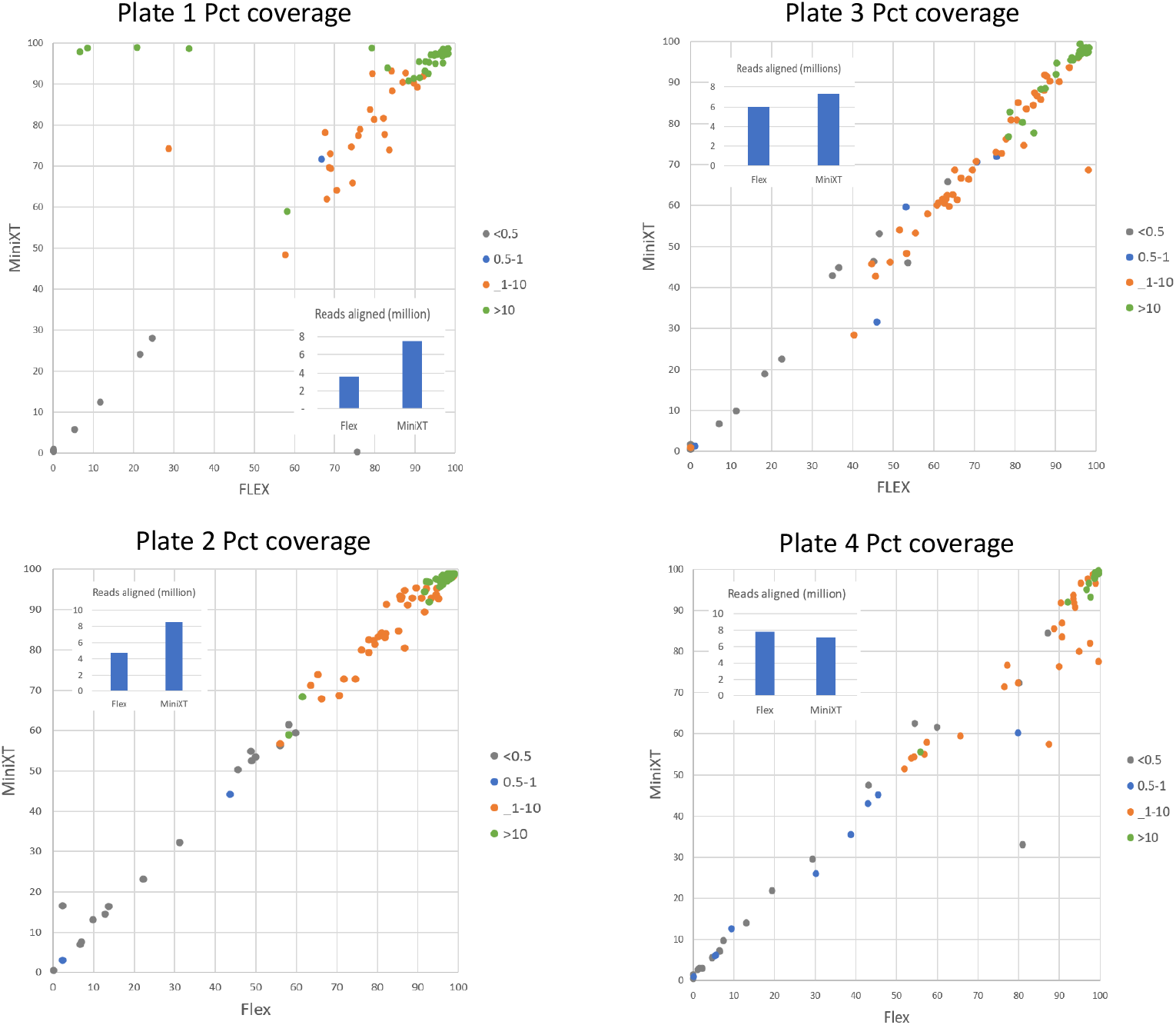
Comparison of individual 96 well plates of samples prepared using the DNA Flex library kit with the same samples prepared with the Mini-XT protocol. The concentrations of the amplicon pools are indicated in ng/ul and broadly correlate with the percentage coverage achieved. The insets indicate the total number of aligned reads for each set of samples.

**Supplementary Figure 4.**
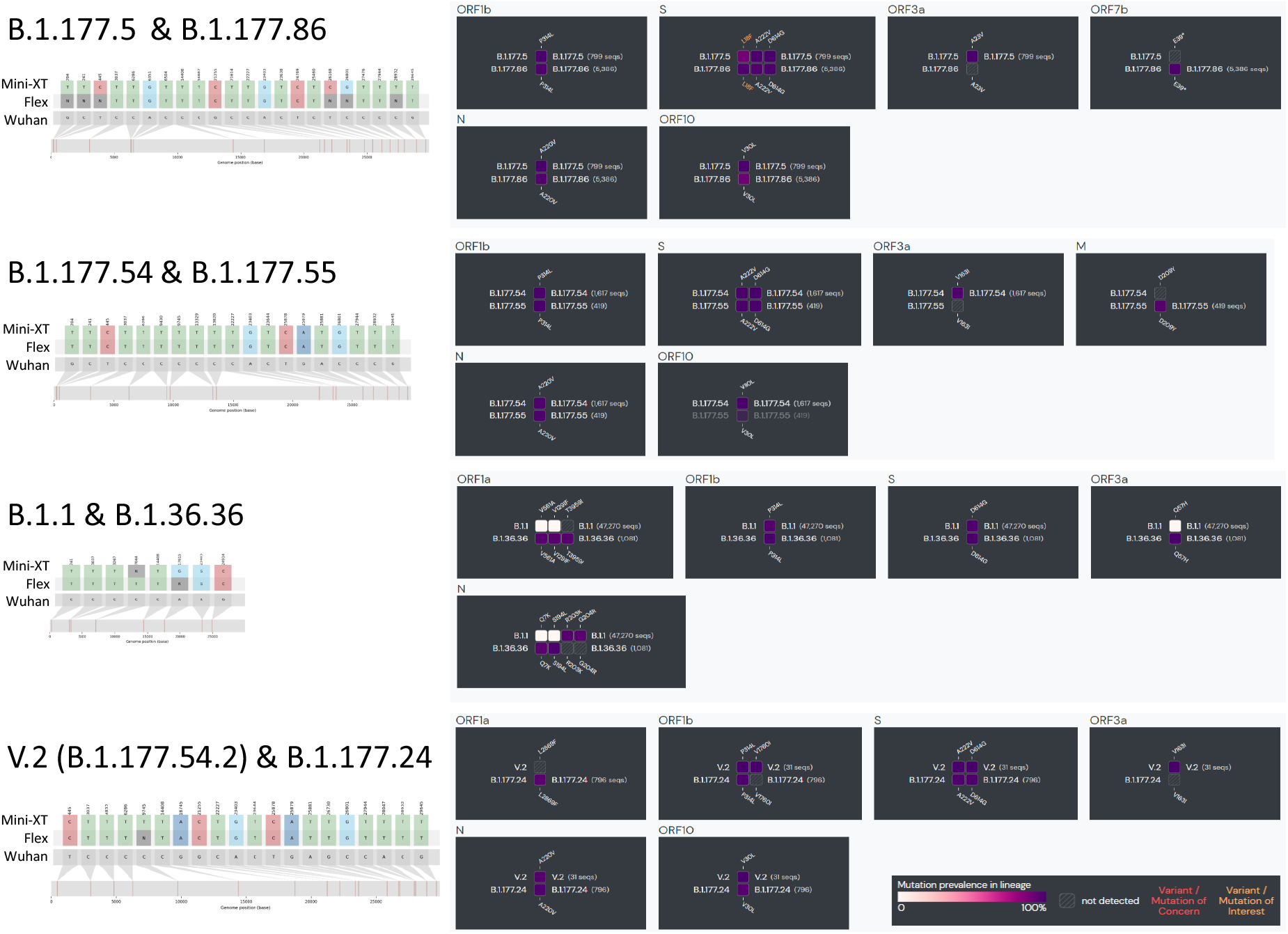
Characteristic variants of sublineages called differentially between sequencing protocols. SNP-IT plots indicate that all the SNPs called from Mini-XT and Flex sequences are either shared or not called in one sequence. The comparison illustrates that most characteristic variants of the discrepant lineages are shared and only vary at several positions.

**Supplementary Figure 5.**
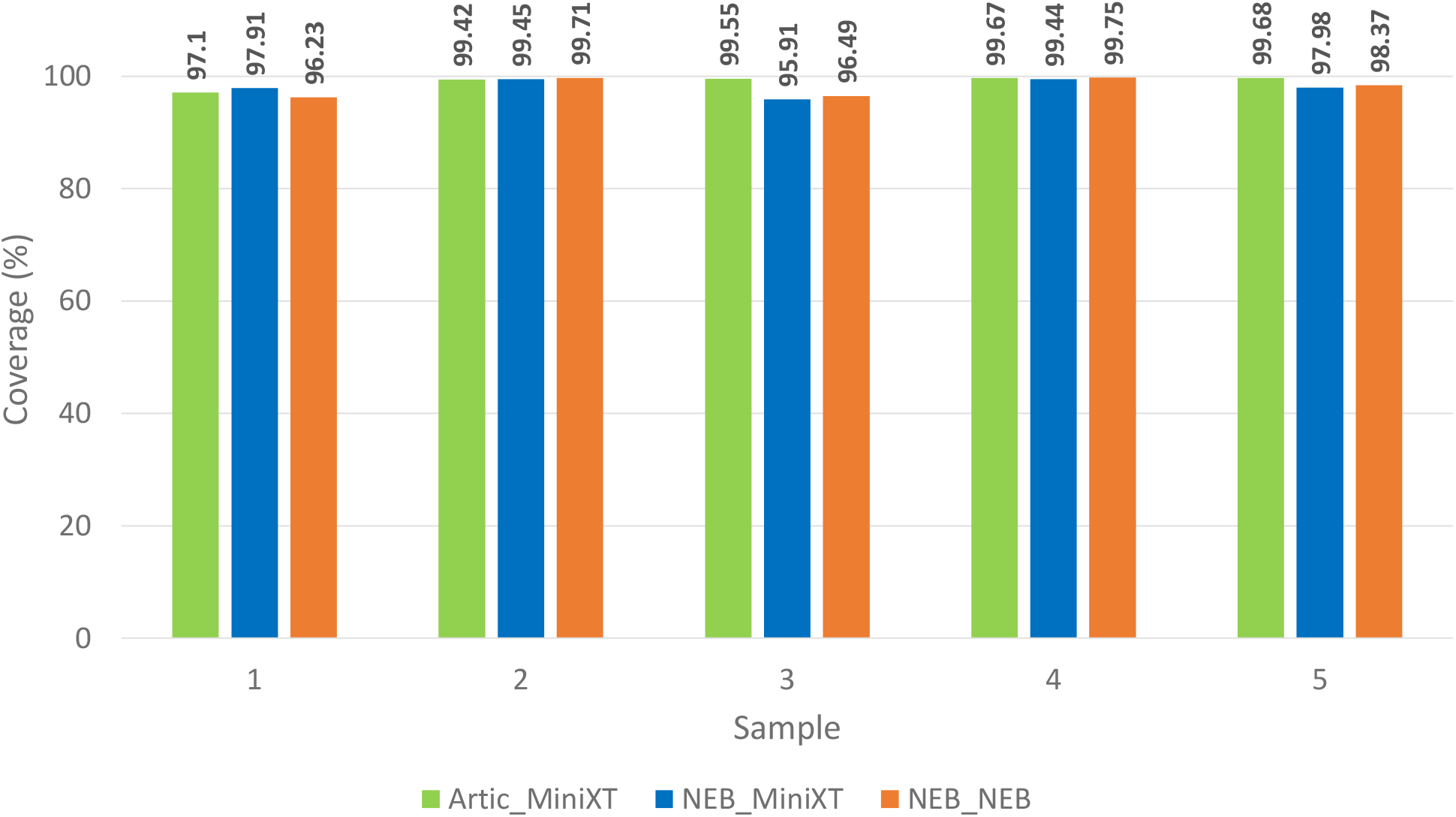
Comparison of Arctic v3 and NEB primer pools. Samples are named according to the amplification and then library preparation method and ordered according to percentage coverage achieved with Articv3 primers and mini-XT library preparation (Artic_MiniXT).

